# Guideline-based Chinese herbal medicine treatment plus standard care for severe coronavirus disease 2019 (G-CHAMPS): evidence from China

**DOI:** 10.1101/2020.03.27.20044974

**Authors:** Yong-an Ye, on behalf of the G-CHAMPS collaborative group

## Abstract

**Objectives:** To assess outcomes in patients who have severe coronavirus disease 2019 (COVID-19) and were treated with either China guideline based Chinese herbal medicines (CHMs) plus standard care or standard care alone.

**Design:** A pilot randomized controlled trial.

**Setting:** Hubei Provincial Hospital of Integrated Chinese and Western Medicine, Wuhan, China

**Patients:** A total of 42 adults with severe COVID-19.

**Interventions:** Participants in the CHM plus standard care group received CHM and standard care, and the control group received standard care alone.

**Measurements and Main Results:** The primary outcome was the change in the disease severity category of COVID-19 after treatment at 7 days. Among 42 participants who were randomized (mean [SD] age 60.43 years [12.69 years]; 21 [50%] were aged ≥ 65 years; and 35 [83%] women, 42 (100%) had data available for the primary outcome. For the primary outcome, one patient from each group died during treatment; the odds of a shift towards death was lower in the CHM plus group than the standard care alone group (common OR 0.59, 95% CI 0.148 to 2.352 *P*=.454). Three (2 from the CHM plus group and 1 from the standard care alone group) patients progressed from severe to critical illness. After treatment, mild, moderate, and severe COVID-19 disease accounted for 18% (5) vs 14% (2), 71% (20) vs 64% (9), and 0% (0) vs 7% (1) of the patients treated with CHM plus standard care vs. standard care alone.

**Conclusions:** For the first time, the G-CHAMPS trial provided valuable information for the national guideline-based CHM treatment for hospitalized patients with severe COVID-19. CHM effects in COVID-19 may be clinically important and warrant further consideration and studies.

## Introduction

Approximately 14-16% patients with Coronavirus disease 2019 (COVID-19) suffer from severe diseases like pneumonia and 5% become critically ill(1, 2). The mortality rate of COVID-19 among those suffering critical illness was reported to be over 50%(1).The National Health Commission and the National Administration of Traditional Chinese Medicine of the People’s Republic of China developed clinical guidelines for the management of COVID-19 (NHC-NATCM-China guidelines)(3, 4). In these guidelines, CHM was included as part of the treatment plans for severe COVID-19. These recommendations were developed by the consensus of experts. We thus conducted this pilot randomized clinical trial (RCT) to test the potential effectiveness of the guideline-based CHM treatment for severe COVID-19 in Wuhan, China.

## Methods

### Study design

This was an open-label, pilot, randomized trial for severe COVID-19. The trial was approved by the ethics committee at Dongzhimen Hospital (No. DZMEC-KY-2020-09). The trial was registered at the Chinese Clinical Trial Registry (ChiCTR2000029418). The trial protocol and protocol amendments were provided in Appendix 1 protocol.

### Patient Enrollment

Patients were screened for eligibility of the G-CHAMPS trial upon admission. During the ongoing epidemic of COVID-19 in Wuhan, China, patients with a confirmatory diagnosis of COVID-19 were directly admitted or transferred to designated COVID-19 hospitals. By Jan 27, 2020, the Chinese government had designated over 40 hospitals for the treatment of COVID-19 in Wuhan. Hubei Provincial Hospital of Integrated Chinese and Western Medicine is one of the government-designated hospitals for the treatment of COVID-19. Inclusion criteria comprised of: adult patients (≥ 18 years), positive test results for SARS-CoV-2 on a polymerase-chain-reaction (PCR) assay, respiratory rate (RR) ≥ 30/min or SaO_2_≤93% or a PaO_2_/FiO_2_ ratio ≤300mmHg(4), and able to provide informed consent. Patients were excluded if known life expectancy was 48 hours or less, on home oxygen at baseline, pregnant or lactating, diagnosed with end stage diseases, or using immunosuppressants for six months or longer. Eligible patients were provided with oral information about the trial and given the opportunity to ask questions. Patients who were willing to take part in the trial were invited for an interview to gather necessary information including a verbal consent; the audio of the interview was electronically recorded.

### Randomization and masking

Eligible participants were randomized with a 2:1 ratio to the CHM plus standard care (CHM plus) group or the standard care alone group, using a simple random allocation method. Allocation was concealed to laboratory personnel and outcome assessors.

### Procedures

Per NHC-NATCM-China guidelines, all patients received standard care, which included hemodynamic monitoring, laboratory testing, supplementary oxygen, intravenous fluids, and routine pharmaceutical medications and other medical care when deemed appropriate by on-duty physicians. Oral ribavirin/arbidole (not remdesivir) was part of the standard care in China (Appendix 1 protocol). Per the NHC-NATCM-China guidelines, patients in the CHM plus group also received CHM within 12 hours after randomization (Appendix 1 protocol). The herbal formulas were supported by Jiangyin Tianjiang Pharmaceutical Co., Ltd. Quality of the herbs was in accordance with the 2015 Chinese Pharmacopoeia(5). All herbs were tested for heavy metals, microbial contamination, and residual pesticides to ensure they meet the safety standards in China prior to use. Trained and experienced technicians prepared the decoction from the formulas according to a standardized procedure; each unit of formula yielded 400mL of decoction, divided into two equal portions. Nurses administered the decoction 200mL to patients orally (via feeding tube if needed) twice daily for a total of seven days in the CHM plus group. Data were retrieved from electronic medical records using the standardized case record forms which created by members of ISARIC(6) (International Severe Acute Respiratory and Emerging Infection Consortium) in collaboration with the World Health Organization.

### Outcomes

The primary outcome was the change in the disease severity category of COVID-19 after treatment. The severity of COVID-19 was assessed based on the Six-Point Clinical Status Scale for COVID-19 (COVID-19 severity scale) (**Score 0**: Hospital discharge or meet discharge criteria-Discharge criteria are defined as: 1 Normal body temperature for more than 3 days; 2 Significantly improved respiratory symptoms: no oxygen supplementation requirement, stable and normal vital signs for longer than 1 day; 3 Lung imaging shows obvious absorption and recsolution of acute infiltrates; 4 Negative results of the nucleic acid test for SARS-CoV-2 for consecutive two times with at least 1 day interval between tests.). (**Score 1**: Mild-Improving and/or mild clinical symptoms and no pneumonia changes in radiological imaging studies.). (**Score 2**: Moderate-Active symptoms like fever and respiratory tract symptoms and pulmonary infiltrates seen in imaging.). (**Score 3**: Severe Meeting any of the following: 1 Respiratory distress, RR ≥30 breaths/min; 2 Pulse oximetry (SpO2) ≤ 93% on room air at rest state; 3 Arterial partial pressure of oxygen (PaO2) / oxygen concentration (FiO2) ≤ 300 mmHg).(**Score 4**: Critical illness Meeting any of the following: 1 Mechanical ventilation; 2 Shock; 3 Other organ failure complications that require intensive care unit care). (**Score 5**: Death). The Six-Point Clinical Status Scale for COVID-19 was defined according to NHC-NATCM-China guideline and WHO R&D Blueprint. An independent clinical event adjudication committee (CEAC) performed the final the outcome assessment based on the pre-specified criteria. Secondary outcomes included the overall survival through last day of treatment, the proportion of patients without improvement (scored 3 to 5 on the COVID-19 severity scale) and the change in serum procalcitonin level after treatment and the prevalence of antibiotic use during treatment.

### Statistical analysis

Since this is a pilot randomized trial, sample size calculation was not performed. For pharmaceutical interventions, a minimum sample size of 12 per group was usually recommended as a rule of thumb for a pilot study(7). Considering a dropout rate of 10%, we aimed to recruit a total sample size of 42 patients (standard care group, n=14; CHM plus group, n=28).

We compared the severity of COVID-19 with ordinal logistic regression (shift analysis). The proportion of patients without clinical improvement after treatment was assessed using the generalized linear model. laboratory findings were performed using the Wilcoxon rank-sum test. Hodges–Lehmann estimates of location shift and 95% CIs were presented.

All outcomes were assessed in the intention-to-treat population with no imputation for missing data. All statistical analyses were performed using SAS version 9.4 (SAS Institute Inc) with a 2-sided p value of less than .05 considered significant.

## Results

Forty-two out of 100 screened patients were included in the trial (Appendix Figure 1). The two groups were generally well balanced at baseline, although older and more women were enrolled in the CHM plus group than the standard care alone group (Table 1). During the G-CHAMPS trial, supportive measures of standard care were similar in the two groups (Appendix 1 protocol).

**Table 1.** **Baseline Demographic and Clinical Characteristics of the Trial Population**.

|  | CHM plus standard care<br>(n=28) | Standard care<br>(n=14) |
| --- | --- | --- |
| **Characteristics** |  |  |
| Age, yr | 65 (53.5-69) | 59 (47-67) |
| Age ≥65 yr, - no. (%) | 16 (57) | 5 (36) |
| Age <65 yr, -no. (%) | 12 (42.863) | 9 (64) |
| Sex, no. (%) |  |  |
| Men | 2 (7) | 4 (29) |
| Women | 25 (93) | 10 (71) |
| Current smoker, no. (%) | 0 | 0 |
| Heart rate, per min | 89 (70-92.5) | 97 (90-105) |
| Blood pressure, -mm Hg |  |  |
| Systolic pressure, mm Hg | 129 (110-140) | 115.5 (110-119) |
| Diastolic pressure, mm Hg | 85 (74.5-90) | 80.5 (75-90) |
| Body temperature, °C | 37 (36.6-37.1) | 36.4 (36.2-37) |
| Respiratory rate >24 breaths, per min | 28 (100) | 14 (100) |
| SaO<sub>2</sub> | 89 (86-90.5) | 89 (87-90) |
| Transfer from other hospitals-no. (%) | 2 (7.41) | 4 (28.57) |
| Onset of symptoms to hospital admission, days | 9 (6.5-11.5) | 9.5 (6-14) |
| Hospital admission to randomization, days | 1 (0.5-2) | 0.5 (0-1) |
| **Any comorbidity-no. (%)** |  |  |
| Chronic heart disease, including congenital heart disease (except hypertension) | 8 (28.57) | 3 (21) |
| Chronic lung disease (except asthma) | 2 (7.14) | 2 (14) |
| Asthma | 1 (3.57) | 0 |
| Mild liver disease | 3 (10.71) | 2 (14) |
| Chronic nervous system diseases | 2 (7.14) | 0 |
| Malignant tumor | 0 | 1 (7.14) |
| Diabetes without complications | 1 (3.57) | 3 (21.43) |
| Hypertension | 12 (42.86) | 7 (50.00) |
| hyperthyroidism | 0 | 1 (7.14) |
| **Presenting symptoms and signs-no. (%)** |  |  |
| Fever* | 27 (96) | 9 (75) |
| Cough | 23 (82) | 12 (86) |
| Sputum | 10 (36) | 4 (29) |
| Sore throat | 1 (4) | 0 |
| Rhinorrhea | 0 | 1 (7) |
| Loss of appetite | 25 (89) | 12 (86) |
| Insomnia | 20 (71) | 10 (71) |
| Wheezing | 5 (18) | 1 (7) |
| Chest pain | 2 (7) | 1 (7) |
| Muscle pain | 8 (29) | 6 (43) |
| Arthralgia | 0 | 1 (7) |
| Fatigue | 26 (93) | 14 (100) |
| Shortness of breath (dyspnea) | 5 (18) | 5 (36) |
| Headache | 2 (7) | 1 (7) |
| Vomiting / nausea | 6 (21) | 1 (7) |
| Diarrhea | 3 (11) | 3 (21) |
| **Chest x-ray and CT findings\*\*** |  |  |
| Ground-glass opacity | 15 (79) | 7 (78) |
| Local patchy shadowing | 0 | 1 (11) |
| Bilateral patchy shadowing | 4 (21) | 1 (11) |

CHM= Chinese herbal medicine. Data are presented as median (IQR) unless otherwise indicated. \*Two participants in the standard care group had no baseline record of fever. \*\* Chest x-ray and CT findings (standard of care plus GC, n=19; standard care group, n=9). Transfer here was considered as new admission in this trial.

**Figure 1.**
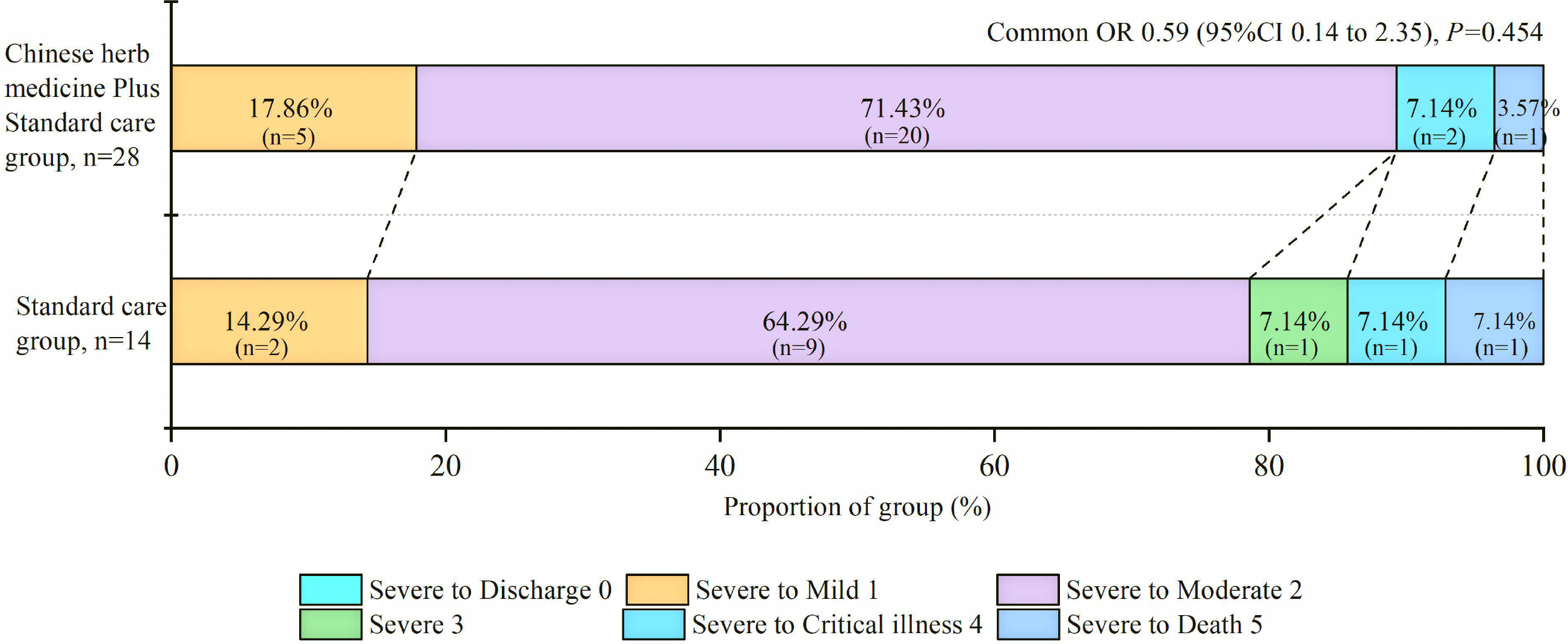
Distribution of COVID-19 severity score at 7 days. OR=odds ratio The figure denotes scores on the COVID-19 severity scale for patients in the Chinese herbal medicine plus standard care group and the standard care alone group. Scores on the COVID-19 severity scale range from 0=discharge to 5=death. A difference between the Chinese herbal medicine plus standard care group and the standard care group was noted in the overall distribution of scores, favoring the Chinese herbal medicine plus standard care group (common odds ratio for improvement of 1 point on the COVID-19, 0.59; 95% confidence interval (CI), 0.14 to 2.35).

For the primary outcome, one patient from each group died during the first three days of treatment; the odds of a shift towards death was lower in the CHM plus group than the standard care group (common OR 0.589, 95% CI 0.148 to 2.352 *P*=.454; Figure 1). For secondary outcomes, 11% (3/28) of patients in the CHM plus group and 21% (3/14) of patients in the standard care alone group had no clinical improvement (difference -10.71 (-35.07 to 13.64), *P*=0.3496) after treatment. More secondary outcomes and safety outcomes were provided in Appendix Table 1-5.

## Discussion

To our best knowledge, this is the first prospective randomized trial to investigate the effect of NHC-NATCM-China guideline-based CHM in patients with severe COVID-19. In this trial, the odds of a shift towards death or critically ill at 7 days after treatment was lower in the CHM plus group, at a non-significant level. The result was collaborated with the universal normalization or near normalization of leukocytes and different inflammatory markers. In a retrospective study with data of 1,099 patients with COVID-19, 5% of the patients were admitted to the ICU, 2% underwent invasive mechanical ventilation, and 1% died; whereas the composite of these endpoints occurred in 25% of the patients with severe disease(8). In our trial, 12% of the patients with severe COVID-19 required ICU care and 5% died within 7 days. Disease severity is an important factor when considering treatment for COVID-19 and likely contributed to the differences between these two studies. An ongoing trial of Gilead Sciences’ Remdesivir utilized a category ordinal scale to define its primary outcome (NCT04257656).

Although COVID-19 is caused by a virus and will heal without treatment in the majority of patients, most patients in the G-CHAMPS trial received antibiotics. The percentages of antibiotic use are comparable to the previous study (80%)(8).

Our study has several limitations, including an open-label design, a small sample size. Additionally, this study lacks long term outcomes and the COVID-19 disease severity scale deserves further investigation. Despite these substantial limitations, the G-CHAMPS trial provided an important opportunity to better understand CHM for severe COVID-19.

For the first time, the G-CHAMPS trial provided valuable information for the national guideline-based CHM treatment for hospitalized patients with severe COVID-19. As effective antiviral treatment is still lacking for COVID-19 and SARS-CoV-2 continues to spread outside of China(9), all potentially effective treatments including CHM worth vigorous further investigation.

## Data Availability

The data that support the findings of this study are available from the corresponding author on reasonable request. Participant data without names and identifiers will be made available after approval from the corresponding author and National Health Commission. After publication of study findings, the data will be available for others to request. The research team will provide an email address for communication once the data are approved to be shared with others. The proposal with detailed description of study objectives and statistical analysis plan will be needed for evaluation of the reasonability to request for our data. The corresponding author and National Health Commission will make a decision based on these materials. Additional materials may also be required during the process.

## Acknowledgments

We thank the International Severe Acute Respiratory and Emerging Infections Consortium (ISARIC) for sharing data collection templates publicly on the website; We thank all patients involved in the study.

## Members of the G-CHAMPS Collaborative Group

In addition to the core writing group, the group members also contributed substantively to the conduct of the G-CHAMPS trial. These authors contributed equally to this work (Appendix 3 the G-CHAMPS collaborative group). All the authors dedicated large amounts of time to the study, in the hope of improving care for patients during COVID-19 outbreak. All members read and approved the final report. All authors agree, as the G-CHAMPS group members, to submit this article.

## References

1. Yang X, Yu Y, Xu J, Shu H, et al: Clinical course and outcomes of critically ill patients with SARS-CoV-2 pneumonia in Wuhan, China: a single-centered, retrospective, observational study. The Lancet Respiratory medicine 2020; February 24.

2. World Health Organization: Coronavirus disease 2019 (COVID-19) Situation Report 41. 2020

3. The National Health Commission and the National Administration of Traditional Chinese Medicine of the People’s Republic of China: Guidance for Corona Virus Disease 2019: Prevention, Control, Diagnosis and Management (Tentative 3rd edition). 2020

4. The National Health Commission and the National Administration of Traditional Chinese Medicine of the People’s Republic of China: Guidance for Corona Virus Disease 2019: Prevention, Control, Diagnosis and Management (Tentative 4th edition). 2020

5. Chinese Pharmacopoeia Commission (2015): Chinese Pharmacopoeia (2015). Beijing:China Medical Science Press, 2015

6. International Severe Acute Respiratory and Emerging Infection Consortium: COVID-19 CRF. 2020

7. Julious SA: Sample size of 12 per group rule of thumb for a pilot study. Pharmaceutical Statistics 2005; 4(4):287–291

8. Guan WJ, Ni ZY, Hu Y, Liang WH, et al: Clinical Characteristics of Coronavirus Disease 2019 in China. The New England journal of medicine 2020; February 28.

9. Wu Z, McGoogan JM: Characteristics of and Important Lessons From the Coronavirus Disease 2019 (COVID-19) Outbreak in China: Summary of a Report of 72314 Cases From the Chinese Center for Disease Control and Prevention. Jama 2020; February 24.

